# Lack of association between *G6PD* variants and Parkinson’s disease

**DOI:** 10.1101/2025.04.23.25324774

**Authors:** Leah V. Chifamba, Sitki Cem Parlar, Lang Liu, Leonard L. Sokol, Eric Yu, Farnaz Asayesh, Jamil Ahmad, Jennifer A. Ruskey, Dan Spiegelman, Cheryl Waters, Oury Monchi, Yves Dauvilliers, Nicolas Dupré, Alla Timofeeva, Anton Emelyanov, Sofya Pchelina, Irina Miliukhina, Lior Greenbaum, Sharon Hassin-Baer, Roy N. Alcalay, Alberto J. Espay, Ziv Gan-Or, Konstantin Senkevich

## Abstract

Oxidative stress has been implicated in Parkinson’s disease (PD). Genes involved in PD, such as *PRKN, PINK1* and *PARK7*, contribute to oxidative stress in dopaminergic neurons. The X-linked G6PD gene encodes glucose 6-phosphate dehydrogenase, an important regulator of oxidative stress. Recent studies suggested that alpha-synuclein aggregates may impair G6PD activity and contribute to dopaminergic neuron loss, and that *G6PD* mutations may independently increase the risk of PD. In this study, we aimed to examine the role of common and rare *G6PD* variants in PD across six cohorts including 8,905 PD patients, 16,770 proxy-patients, 394,098 controls. These cohorts were analyzed after stratification by sex and then combined to account for the *G6PD* X-linked location. Using logistic regression, we did not identify significant associations for common variants in any of the cohorts. The Combined and Multivariate Collapsing Wald (CMC-Wald) test was performed to assess the cumulative effect of rare variants (minor allele frequency < 0.01) across six cohorts, followed by a meta-analysis, also demonstrating lack of association. In conclusion, we did not find evidence for a role for *G6PD* in PD.

## 1. Introduction

Oxidative stress is one of the factors playing a role in the pathogenesis of Parkinson’s disease (PD) (Abraham et al. 2005; Dias, Junn, and Mouradian 2013; Hauser and Hastings 2013). Core genes implicated in the PD pathway, such as *PRKN, PINK1* and *PARK7*, contribute to oxidative stress, impacting dopaminergic neurons (Dorszewska et al. 2020; Chang and Chen 2020).

Glucose 6-phosphate dehydrogenase (*G6PD*) is an X-linked gene that encodes for the G6PD enzyme, which regulates oxidative stress (Stanton 2012; Tang 2019). The enzyme is a critical component of the pentose phosphate pathway, where it catalyzes the production of nicotinamide adenine dinucleotide phosphate (NADPH), thereby preventing cellular damage (Efferth et al. 2006). The role of G6PD has mostly been investigated in hemolytic anemia, where G6PD deficiency leads to oxidative damage in erythrocytes (Luzzatto, Ally, and Notaro 2020). While G6PD deficiency predominantly affects males (Domingo et al. 2019), it has also manifested in females carrying two mutated copies of the gene (Bain et al. 2023; Rameli et al. 2023; Boussaadni et al. 2022). G6PD deficiency is particularly prevalent among Jewish populations of Middle Eastern and North African descent (Alangari et al. 2023).

A recent study suggested that alpha-synuclein aggregates, a feature of most PD patients, may lead to loss of G6PD within synaptic vesicles, resulting in decreased NADPH and oxidative damage in dopaminergic neurons (Stykel et al. 2025). The authors also suggested a genetic association between *G6PD* missense mutations and PD (Stykel et al. 2025). *G6PD* has not been identified as associated with PD in previous X-wide association studies, although it is located closely to an associated locus (Le Guen et al. 2021; Leal et al. 2023). Furthermore, another study has shown that deletion of *G6PD* using CRISPR-Cas9 impacted PINK1-Parkin-mediated mitophagy (Cho et al. 2024), a pathway involved in PD (Vizziello et al. 2021). This suggests that deficiency in G6PD may intensify mitochondrial dysfunction and oxidative stress, potentially contributing to the pathogenesis of PD.

Our study aimed to gain further understanding of the involvement of *G6PD* in PD by investigating the potential role of common and rare *G6PD* genetic variants in PD. The analysis included six independent cohorts with a total of 8,905 PD cases, 16,770 proxy-cases, 394,098 controls. The cohorts were analyzed after stratifying by sex and then combined to account for the *G6PD* X-linked location.

## 2. Methods

### 2.1 Study participants

The population for the genetic analysis comprised six cohorts, 8,905 PD cases, 16,770 proxy-cases, 394,098 controls (Table 1). The first four cohorts were collected at McGill University and have been previously reported. In brief, they include i) French-Canadian/French cohort collected from Quebec, Canada (Gan-Or et al. 2020) and Montpellier, France, ii) Columbia University cohort, New York (Alcalay et al. 2016), iii) Sheba cohort from Sheba Medical Center, Israel (Ruskey et al. 2019). This cohort comprised Ashkenazi and Mizrahi Jews iv) a cohort from Pavlov First State Medical University and Institute of Human Brain, Russia (Senkevich et al. 2023). Additional cohorts included: (i) Whole-genome sequencing (WGS) data from Accelerated Medicines Partnership - Parkinson’s Disease (AMP-PD, https://amp-pd.org/, data assessed using the Terra platform), (ii) The UK Biobank (UKBB) including proxy-patients (first-degree relatives of PD patients) was acquired using WGS data from the UK Biobank Research Analysis Platform (https://www.ukbiobank.ac.uk/). Ethics approval for the research study was granted by the McGill University Research Ethics Board.

**Table 1:** Demographics of studied cohorts.

|  |  |  |  |  |  | Mean age (SD) | Mean age (SD) |
| --- | --- | --- | --- | --- | --- | --- | --- |
| Cohort | N,<br>Cases | N,<br>controls | Sex | N,<br>Cases | N,<br>Controls | N, Cases | N, Controls |
| McGill University | 1,027 | 1,115 | M<br>F | 636<br>391 | 517<br>598 | 59.71 (11.41)<br>60.39 (11.46) | 54.43 (14.33)<br>54.25 (13.47) |
| Columbia | 1,070 | 492 | M<br>F | 691<br>379 | 172<br>320 | 59.38 (11.65)<br>60.09 (11.89) | 67.11 (11.17)<br>62.38 (10.04) |
| Pavlov and Human<br>Brain Institute | 469 | 332 | M<br>F | 182<br>287 | 113<br>219 | 61.40 (16.74)<br>62.56 (17.41) | 68.48 (21.81)<br>75.33 (14.58) |
| Sheba | 984 | 525 | M<br>F | 603<br>381 | 296<br>229 | 63.82 (12.58)<br>64.58 (13.11) | 33.45 (8.93)<br>33.31 (6.69) |
| AMP-PD | 2,341 | 3,486 | M<br>F | 1,459<br>882 | 1,648<br>1,838 | 64.80 (9.82)<br>63.29 (10.04) | 69.04 (13.58)<br>67.73 (13.45) |
| UKBB (with proxies) | 19,736 | 387,955 | M<br>F | 9,007<br>10,729 | 178,331<br>209,624 | 59.57 (7.72)<br>58.90 (7.65) | 57.03 (7.55)<br>56.61 (7.52) |

Key: N, Number; M, Male; F, Female; SD, standard deviation; UKBB, UK biobank; AMP-PD, Accelerating Medicines Partnership Parkinson's disease

### 2.2 G6PD sequencing and quality control

Targeted next-generation sequencing of *G6PD* was performed using molecular inversion probes (MIPs) in the four cohorts gathered at McGill University as previously described (Rudakou et al. 2020). The MIPs protocol is accessible at https://github.com/gan-orlab/MIP_protocol. The Genome Quebec Innovation Centre carried out the sequencing utilizing the Illumina NovaSeq 6000 SP PE100 platform. Burrows-Wheeler Aligner (hg19) was used for alignment (Li and Durbin 2009), and Genome Analysis Toolkit (GATK, v3.8) was employed for post-alignment quality control and variant calling (McKenna et al. 2010). Using PLINK program v1.9 (Purcell et al. 2007), we carried out quality control by eliminating variants and samples of lower quality. SNPs with missingness of more than 10% were excluded from the analysis. Variants having a minimum quality score (GQ) of 30 and minimal depths of coverage 30x were included.

As previously described, quality control procedures for WGS for AMP-PD cohorts were carried out on an individual and variant level (https://amp-pd.org/whole-genome-data) (Iwaki et al. 2021). We performed quality control on the UKBB WGS data using the Genome Analysis Toolkit (GATK, v3.8), using a minimum depth of coverage of 30x and GQ of 20 for further analysis. Hg38 reference was applied for AMP-PD and UKBB.

### 2.3 Statistical Analyses

Since *G6PD* is located on the X-chromosome, we stratified the cohorts by sex to account for differences in allele dosage, analyzing males and females separately. The results were then meta-analyzed across cohorts. To assess the association between common variants (minor allele frequency, MAF > 0.01) in *G6PD* with PD, we conducted logistic regression adjusting for age, using PLINK v1.9 (Purcell et al. 2007). We also used the Combined and Multivariate Collapsing (CMC) Wald burden test to study the association of rare variants (MAF < 0.01) with PD, and performed a meta-analysis using the metagen package (Shim and Kim 2019). We examined in each cohort the burden of five groups of variants: i) all rare variants, ii) nonsynonymous variants, iii) functional variants (including stop/frameshift, splicing and nonsynonymous variants), iv) variants with a high Combined Annotation Dependent Deletion (CADD) score ≥20 and v) loss-of-function variants. Additionally, we included *G6PD* variants associated with mean enzyme activity of less than 20% of normal (Nannelli et al. 2023). These variants were selected based on the 2024 WHO classification of *G6PD* variants, specifically Class A, which includes variants that significantly reduce enzyme activity and are associated with chronic hemolytic anemia (Luzzatto et al. 2024). The description of these variants is summarized in Supplementary Table 1. For multiple testing correction, we applied Bonferroni correction for common variant analysis and the False Discovery Rate (FDR) method using Benjamini-Hochberg for CMC-Wald burden test.

## 3. Results

We sequenced *G6PD* in four cohorts at McGill, achieving an average read depth of 1068x with >89% nucleotides covered at >30x. We discovered and analyzed 9 common variants (MAF>1%) from all cohorts for their association with PD, none of which remained significant after Bonferroni correction (Supplementary Table 2). We found 112 rare variants with MAF <1% in cohorts sequenced at McGill, 82 in AMP-PD and 6,106 in UKBB cohort (Supplementary Table 3). Burden analysis before and after sex stratification, using CMC-Wald, showed no association after meta-analysis in any of the variant categories including variants associated with reduced G6PD activity. Individual cohort results are summarized in Supplementary Table 4.

**Table 2:** Meta-analysis results of CMC burden test of six *G6PD* variants^i^ previously linked to PD.

|  | Effect size<br>estimate | 95% confidence<br>interval | Z-statistic | P-value |
| --- | --- | --- | --- | --- |
| Meta-analysis - all | 0.0810 | [-0.2431; 0.4052] | 0.49 | 0.306 |
| Meta-analysis - males | 0.2807 | [-0.2420; 0.8034] | 1.05 | 0.293 |
| Meta-analysis - females | 0.0135 | [-0.6348; 0.6617] | 0.04 | 0.968 |

<sup>i</sup>Six *G6PD* variants are: rs137852318, X:154533067:A:T (hg38), X:154534437:G:A (hg38), rs1557229675, rs1050829, rs5030870

We also analyzed the allele frequencies of the six rare *G6PD* missense variants reported by Stykel et al., (Stykel et al. 2025) in our cohorts (Supplementary Table 5). Using CMC-Wald test, we assessed these variants both combined and stratified by sex and did not find significant association after meta-analysis (Table 2).

## 4. Discussion

This study aimed to assess the association of common and rare *G6PD* variants with PD in six independent cohorts. None of the associations remained significant after multiple correction and did not reach significance in the meta-analysis. Overall, our findings suggest lack of genetic associations between common and rare *G6PD* variants with PD.

A recent study (Stykel et al. 2025), showed that alpha-synuclein aggregates may impair G6PD activity, contributing to dopamine loss, using UKBB data from GeneBass (Karczewski et al. 2022), they suggested that *G6PD* variants may independently increase the risk of PD. However, our analysis of six independent cohorts does not support this association. We also utilized UKBB data as one of the cohorts and included proxy-cases, which may have resulted in different allele frequencies compared to their study. Additionally, we conducted an in-depth analysis stratified by sex to account for the X-chromosome location of *G6PD*, an approach that was not implemented in the GeneBass study. Despite all our efforts, we did not find compelling genetic evidence for the involvement of this gene in PD.

To the best of our knowledge, parkinsonism has not been previously described in patients with G6PD deficiency, making a direct causative link between G6PD deficiency and PD unlikely. Some medications, including levodopa, have been identified to exacerbate anemia symptoms in patients with G6PD deficiency (Richardson and O’Malley 2025). Thus, current clinical evidence does not support a role for G6PD deficiency in PD.

Our study has several limitations. We used the CMC-Wald burden test, which collapses all rare variants and assumes a unidirectional effect on the trait, making it less robust. However, this approach allows for signal detection, and given the absence of positive results, we did not perform additional kernel association tests. To account for this limitation, we conducted multiple stratifications, including analyses restricted to variants associated with a significant reduction in G6PD enzymatic activity. The study comprised mostly individuals of European ancestry, which limits genetic diversity. Finally, different sequencing methods were used for the data sequenced at McGill compared to WGS data from AMP-PD and UKBB, potentially introducing variability in variant detection across cohorts.

In conclusion, our analyses showed a lack of association between *G6PD* common and rare variants with PD, therefore, future studies should further investigate the role of other oxidative stress related genes in PD.

## Data availability

All the data generated in this study are provided within the manuscript. https://github.com/gan-orlab/G6PD-on-PD

## Acknowledgements

We would like to sincerely thank the participants from the various cohorts who contributed to this study. This research was partially funded by the Canada First Research Excellence Fund through McGill University’s Healthy Brains, Healthy Lives initiative, with additional support from Calcul Québec and Compute Canada. Access to UK Biobank data was accessed using the UKBiobank Research Analysis Platform (https://www.ukbiobank.ac.uk/) under application number 45551. Access to data was supported by the NeuroHub infrastructure. Data for this study was also sourced from the AMP-PD Knowledge Platform. More information about the study can be found at https://www.amp-pd.org. AMP-PD, a public-private partnership managed by the Foundation for the National Institutes of Health (FNIH), is funded by Celgene, GSK, the Michael J. Fox Foundation for Parkinson’s Research, the National Institute of Neurological Disorders and Stroke, Pfizer, AbbVie, Sanofi, and Verily.

Genetic data for this research were obtained from several sources, including the Fox Investigation for New Discovery of Biomarkers (BioFIND), the Harvard Biomarker Study (HBS), the Parkinson’s Progression Markers Initiative (PPMI), the Parkinson’s Disease Biomarkers Program (PDBP), the International LBD Genomics Consortium (iLBDGC), and the STEADY-PD III Investigators. BioFIND is sponsored by The Michael J. Fox Foundation for Parkinson’s Research (MJFF), with support from the National Institute for Neurological Disorders and Stroke (NINDS). The BioFIND Investigators were not involved in the review of data analysis or the manuscript content.

The HBS is a collaborative effort of investigators, with a full list available at https://www.bwhparkinsoncenter.org/biobank/, and is funded by philanthropy, NIH, and non-NIH sources. HBS investigators did not participate in the review of data or manuscript content. PPMI is a public-private partnership funded by the Michael J. Fox Foundation for Parkinson’s Research and its partners, listed at www.ppmi-info.org/fundingpartners. PPMI investigators were also not involved in reviewing data or the manuscript content. More details on the study are available at www.ppmi-info.org. The PDBP consortium is supported by the NINDS at the NIH, and a full list of PDBP investigators can be found at https://pdbp.ninds.nih.gov/policy. The PDBP investigators did not review the data analysis or manuscript content.

The “Genome Sequencing in Lewy Body Dementia and Neurologically Healthy Controls: A Resource for the Research Community” dataset was created by the iLBDGC, co-directed by Dr. Bryan J. Traynor and Dr. Sonja W. Scholz of the NIH Intramural Research Program. The iLBDGC investigators did not participate in reviewing the data analysis or manuscript content. For a full list of contributions, refer to For a full list of contributions, refer to DOI: 10.1038/s41588-021-00785-3. STEADY-PD III is a 36-month, Phase 3, placebo-controlled trial assessing the efficacy of isradipine (10 mg daily) in 336 participants with early-stage Parkinson’s disease. The trial was funded by NINDS and supported by the Michael J. Fox Foundation for Parkinson’s Research and the Parkinson Study Group. The STEADY-PD III investigators did not review the data analysis or manuscript content. A full list of investigators can be found at https://clinicaltrials.gov/ct2/show/NCT02168842. ZGO is supported by the Chercheurs-boursiers award from the Fonds de recherche du Québec – Santé (FRQS) in collaboration with Parkinson Quebec and is a William Dawson Scholar. Access to certain participants for this research was facilitated by the Quebec Parkinson’s Network (http://rpq-qpn.ca/en/).

## CRediT authorship contribution statement

**Leah V. Chifamba:** Conceptualization, Data curation, Formal analysis, Visualization, Writing - original draft, Writing - Review and Editing. **Sitki Cem Parlar:** Data curation, Formal analysis, Software, Writing - review and editing. **Lang Liu:** Data curation, Formal analysis, Software, Writing - review and editing. **Leonard L. Sokol:** Resources, Writing - review and editing. **Eric Yu:** Data curation, Formal analysis, Software, Writing - review and editing. **Farnaz Asayesh:** Investigation, Writing - review and editing. **Jamil Ahmad:** Investigation, Writing - review and editing. **Jennifer A. Ruskey:** Investigation, Methodology, Writing - review and editing. **Dan Spiegelman:** Data curation, Formal analysis, Software, Writing - review and editing. **Cheryl Waters:** Resources, Writing – review and editing. **Oury Monchi:** Resources, Writing - review and editing. **Yves Dauvilliers:** Resources, Writing - review and editing. **Nicolas Dupré:** Resources, Writing - review and editing. **Alla Timofeeva:** Resources, Writing - review and editing. **Anton Emelyanov:** Resources, Writing - review and editing. **Irina Miliukhina:** Resources, Writing - review and editing. **Sofya Pchelina:** Resources, Writing - review and editing. **Lior Greenbaum:** Resources, Writing - review and editing. **Sharon Hassin-Baer:** Resources, Writing - review and editing. **Roy N. Alcalay:** Resources, Writing - review and editing. **Alberto J. Espay:** Resources, Writing - review and editing. **Ziv Gan-Or:** Conceptualization, Funding acquisition, Methodology, Project administration, Resources, Supervision, Visualization, Writing - review and editing. **Konstantin Senkevich:** Conceptualization, Methodology, Supervision, Visualization, Writing - review and editing.

## Abbreviations

AMP-PD: Accelerating Medicines Partnership Parkinson’s disease
UKBB: UK biobank
G6PD: Glucose 6-phosphate dehydrogenase
CMC: Combined and Multivariate Collapsing Wald
CADD: Combined Annotation Dependent Deletion
CADD: Parkinson’s Disease

## References

[1] Abraham, S., C. C. Soundararajan, S. Vivekanandhan, and M. Behari. 2005. “Erythrocyte Antioxidant Enzymes in Parkinson’s Disease.” The Indian Journal of Medical Research 121 (2): 111–15.

[2] Alangari, Abdulaziz S., Ashraf A. El-Metwally, Abdullah Alanazi, Badr F. Al Khateeb, Hanan M. Al Kadri, Ibtehaj F. Alshdoukhi, Aljohrah I. Aldubikhi, Muzun Alruwaili, and Awad Alshahrani. 2023. “Epidemiology of Glucose-6-Phosphate Dehydrogenase Deficiency in Arab Countries: Insights from a Systematic Review.” Journal of Clinical Medicine 12 (20): 6648. 10.3390/jcm12206648.

[3] Alcalay, Roy N., Oren A. Levy, Pavlina Wolf, Petra Oliva, Xiaokui Kate Zhang, Cheryl H. Waters, Stanley Fahn, et al. 2016. “SCARB2 Variants and Glucocerebrosidase Activity in Parkinson’s Disease.” Npj Parkinson’s Disease 2 (1): 1–4. 10.1038/npjparkd.2016.4.

[4] Bain, Barbara J., Jane Myburgh, Kirstin Lund, and Aristeidis Chaidos. 2023. “G6PD Deficiency in Patients Identified as Female.” American Journal of Hematology 98 (2): 359–60. 10.1002/ajh.26704.

[5] Boussaadni, Yousra El, Abdelhakim Aboulfouyoul, Kaoutar Khabbache, Hanan Khalki, and Abdallah Oulmaati. 2022. “Glucose-6-Phosphate Dehydrogenase (G6PD) Deficiency in Girls: A Diagnosis Not to Be Missed (a Case Report).” The Pan African Medical Journal 42 (July):240. 10.11604/pamj.2022.42.240.36270.

[6] Chang, Kuo-Hsuan, and Chiung-Mei Chen. 2020. “The Role of Oxidative Stress in Parkinson’s Disease.” Antioxidants 9 (7): 597. 10.3390/antiox9070597.

[7] Cho, Yik-Lam, Hayden Weng Siong Tan, Jicheng Yang, Basil Zheng Mian Kuah, Nicole Si Ying Lim, Naiyang Fu, Boon-Huat Bay, Shuo-Chien Ling, and Han-Ming Shen. 2024. “Glucose-6-Phosphate Dehydrogenase Regulates Mitophagy by Maintaining PINK1 Stability.” Life Metabolism 4 (1): oae040. 10.1093/lifemeta/loae040.

[8] Dias, Vera, Eunsung Junn, and M. Maral Mouradian. 2013. “The Role of Oxidative Stress in Parkinson’s Disease.” Journal of Parkinson’s Disease 3 (4): 461–91. 10.3233/JPD-130230.

[9] Domingo, Gonzalo J., Nicole Advani, Ari W. Satyagraha, Carol H. Sibley, Elizabeth Rowley, Michael Kalnoky, Jessica Cohen, Michael Parker, and Maureen Kelley. 2019. “Addressing the Gender-Knowledge Gap in Glucose-6-Phosphate Dehydrogenase Deficiency: Challenges and Opportunities.” International Health 11 (1): 7. 10.1093/inthealth/ihy060.

[10] Dorszewska, Jolanta, Marta Kowalska, Michał Prendecki Thomas Piekut, Joanna Kozłowska, and Wojciech Kozubski. 2020. “Oxidative Stress Factors in Parkinson’s Disease.” Neural Regeneration Research 16 (7): 1383–91. 10.4103/1673-5374.300980.

[11] Efferth, T., S. M. Schwarzl, J. Smith, and R. Osieka. 2006. “Role of Glucose-6-Phosphate Dehydrogenase for Oxidative Stress and Apoptosis.” Cell Death & Differentiation 13 (3): 527–28. 10.1038/sj.cdd.4401807.

[12] “G6PD Glucose-6-Phosphate Dehydrogenase [Homo Sapiens (Human)] - Gene - NCBI.” n.d. Accessed January 18, 2023. https://www.ncbi.nlm.nih.gov/gene/2539.

[13] Gan-Or, Ziv, Trisha Rao, Etienne Leveille, Clotilde Degroot, Sylvain Chouinard, Francesca Cicchetti, Alain Dagher, et al. 2020. “The Quebec Parkinson Network: A Researcher-Patient Matching Platform and Multimodal Biorepository.” Journal of Parkinson’s Disease 10 (1): 301–13. 10.3233/JPD-191775.

[14] Hauser, David N, and Teresa G Hastings. 2013. “Mitochondrial Dysfunction and Oxidative Stress in Parkinson’s Disease and Monogenic Parkinsonism.” Neurobiology of Disease 51 (March):35–42. 10.1016/j.nbd.2012.10.011.

[15] Iwaki, Hirotaka, Hampton L. Leonard, Mary B. Makarious, Matt Bookman, Barry Landin, David Vismer, Bradford Casey, et al. 2021. “Accelerating Medicines Partnership: Parkinson’s Disease. Genetic Resource.” Movement Disorders 36 (8): 1795–1804. 10.1002/mds.28549.

[16] Karczewski, Konrad J., Matthew Solomonson, Katherine R. Chao, Julia K. Goodrich, Grace Tiao, Wenhan Lu, Bridget M. Riley-Gillis, et al. 2022. “Systematic Single-Variant and Gene-Based Association Testing of Thousands of Phenotypes in 394,841 UK Biobank Exomes.” Cell Genomics 2 (9). 10.1016/j.xgen.2022.100168.

[17] Le Guen, Yann, Valerio Napolioni, Michael E. Belloy, Eric Yu, Lynne Krohn, Jennifer A. Ruskey, Ziv Gan-Or, Gabriel Kennedy, Sarah J. Eger, and Michael D. Greicius. 2021. “Common X-Chromosome Variants Are Associated with Parkinson Disease Risk.” Annals of Neurology 90 (1): 22–34. 10.1002/ana.26051.

[18] Leal, Thiago P., Jennifer N. French-Kwawu, Mateus H. Gouveia, Victor Borda, Miguel Inca-Martinez, Emily A. Mason, Andrea RVR Horimoto, et al. 2023. “X-Chromosome Association Study in Latin American Cohorts Identifies New Loci in Parkinson Disease.” medRxiv. 10.1101/2023.01.31.23285199.

[19] Li, Heng, and Richard Durbin. 2009. “Fast and Accurate Short Read Alignment with Burrows– Wheeler Transform.” Bioinformatics 25 (14): 1754–60. 10.1093/bioinformatics/btp324.

[20] Luzzatto, Lucio, Mwashungi Ally, and Rosario Notaro. 2020. “Glucose-6-Phosphate Dehydrogenase Deficiency.” Blood 136 (11): 1225–40. 10.1182/blood.2019000944.

[21] Luzzatto, Lucio, Germana Bancone, Pierre-Antoine Dugué, Weiying Jiang, Angelo Minucci, Caterina Nannelli, Daniel Pfeffer, et al. 2024. “New WHO Classification of Genetic Variants Causing G6PD Deficiency.” Bulletin of the World Health Organization 102 (8): 615–17. 10.2471/BLT.23.291224.

[22] McKenna, Aaron, Matthew Hanna, Eric Banks, Andrey Sivachenko, Kristian Cibulskis, Andrew Kernytsky, Kiran Garimella, et al. 2010. “The Genome Analysis Toolkit: A MapReduce Framework for Analyzing next-Generation DNA Sequencing Data.” Genome Research 20 (9): 1297–1303. 10.1101/gr.107524.110.

[23] Nannelli, Caterina, Andrea Bosman, Jane Cunningham, Pierre-Antoine Dugué, and Lucio Luzzatto. 2023. “Genetic Variants Causing G6PD Deficiency: Clinical and Biochemical Data Support New WHO Classification.” British Journal of Haematology 202 (5): 1024– 32. 10.1111/bjh.18943.

[24] Purcell, Shaun, Benjamin Neale, Kathe Todd-Brown, Lori Thomas, Manuel A. R. Ferreira, David Bender, Julian Maller, et al. 2007. “PLINK: A Tool Set for Whole-Genome Association and Population-Based Linkage Analyses.” American Journal of Human Genetics 81 (3): 559–75. 10.1086/519795.

[25] Rameli, Nabilah, Justin Hor Hung Juan, Ling Pei Chi, Nurul Asyikin Nizam Akbar, Adibah Daud, Syamihah Mardhiah A. Razak, and Sumaiyah Adzahar. 2023. “Severe Hemolysis Triggered by Favism in a Female Patient with G6PD Deficiency: A Case Report.” Biomedical Research and Therapy 10 (10): 5953–55. 10.15419/bmrat.v10i10.836.

[26] Richardson, S. Russ, and Gerald F. O’Malley. 2025. “Glucose-6-Phosphate Dehydrogenase Deficiency.” In StatPearls. Treasure Island (FL): StatPearls Publishing. http://www.ncbi.nlm.nih.gov/books/NBK470315/.

[27] Rudakou, Uladzislau, Jennifer A. Ruskey, Lynne Krohn, Sandra B. Laurent, Dan Spiegelman, Lior Greenbaum, Gilad Yahalom, et al. 2020. “Analysis of Common and Rare VPS13C Variants in Late-Onset Parkinson Disease.” Neurology: Genetics 6 (1): 385. 10.1212/NXG.0000000000000385.

[28] Ruskey, Jennifer A., Lior Greenbaum, Léanne Roncière, Armaghan Alam, Dan Spiegelman, Christopher Liong, Oren A. Levy, et al. 2019. “Increased Yield of Full GBA Sequencing in Ashkenazi Jews with Parkinson’s Disease.” European Journal of Medical Genetics 62 (1): 65–69. 10.1016/j.ejmg.2018.05.005.

[29] Senkevich, Konstantin, Mariia Beletskaia, Aliza Dworkind, Eric Yu, Jamil Ahmad, Jennifer A. Ruskey, Farnaz Asayesh, et al. 2023. “Association of Rare Variants in ARSA with Parkinson’s Disease.” medRxiv. 10.1101/2023.03.08.23286773.

[30] Shim, Sung Ryul, and Seong-Jang Kim. 2019. “Intervention Meta-Analysis: Application and Practice Using R Software.” Epidemiology and Health 41 (March):e2019008. 10.4178/epih.e2019008.

[31] Stanton, Robert C. 2012. “Glucose-6-Phosphate Dehydrogenase, NADPH, and Cell Survival.” Iubmb Life 64 (5): 362–69. 10.1002/iub.1017.

[32] Stykel, Morgan G., Shehani V. Siripala, Eric Soubeyrand, Carla L. Coackley, Ping Lu, Suelen Camargo, Sharanya Thevasenan, et al. 2025. “G6PD Deficiency Triggers Dopamine Loss and the Initiation of Parkinson’s Disease Pathogenesis.” Cell Reports 44 (1). 10.1016/j.celrep.2024.115178.

[33] Tang, Bor Luen. 2019. “Neuroprotection by Glucose-6-Phosphate Dehydrogenase and the Pentose Phosphate Pathway.” Journal of Cellular Biochemistry 120 (9): 14285–95. 10.1002/jcb.29004.

[34] Vizziello, Maria, Linda Borellini, Giulia Franco, and Gianluca Ardolino. 2021. “Disruption of Mitochondrial Homeostasis: The Role of PINK1 in Parkinson’s Disease.” Cells 10 (11): 3022. 10.3390/cells10113022.

